# A Tabular Residual Neural Network for Diabetes Classification and Prediction

**DOI:** 10.64898/2025.12.29.25343132

**Authors:** Alessandro Hammond, Musab Afridi, Keshav Balakrishna

## Abstract

Diabetes Mellitus (DM) is a metabolic disorder characterized by hyperglycemia, with type 1 characterized as an autoimmune destruction of pancreatic beta cells and type 2 characterized by insulin resistance with progressive beta cell dysfunction. This study applied an existing binary classification algorithm (ALTARN) to accurately predict DM. ALTARN, as a tabular attention residual neural network, uses residual connection to find complex patterns present in tabular columns. We achieved an average training accuracy of 75.22%. Furthermore, a robust set of validation metrics was obtained via five-fold stratified cross-validation, yielding an average accuracy of 74.61%, an average precision of 72.36%, a mean recall of 79.69%, and a mean F1 score of 75.83%.

## I. Introduction

Diabetes Mellitus (DM) is a persistent metabolic disease characterized by hyperglycemia due to pancreatic beta cell dysfunction. As of 2024, approximately 589 million individuals live with diabetes worldwide [2]. As the incidence continues to rise in low-income countries [3]–[5], Artificial Intelligence (AI) and Machine Learning (ML) have been shown to be accurate and robust tools in the binary classifications of disease due to their ability to identify and utilize more complex non-linear patterns in the tabular input data. AI can facilitate earlier diagnosis through pattern recognition in clinical data, which allows clinicians to intervene before complications develop and optimize treatment strategies. Models must retain feature representations from prior layers to decode complex patterns effectively. Recurrent Neural Networks accomplish this through temporal feedback, while Residual Neural Networks use additive skip connections. While Recurrent Neural Networks have been used in the binary classification of DM [6], Residual Neural Networks (ResNets) have not been extensively explored as a standalone approach beyond comparative use, despite work showing they are capable of accurate classification in many field of medicine [1], [7]. As such, ALTARN model adapts a pre-existing algorithm to DM.

## II. METHODOLOGY

To evaluate the effectiveness of ALTARN on diabetes classification, we applied the existing algorithm to a new diabetes dataset. ALTARN (Attention-based Tabular Residual Neural Network) was originally designed for binary classification tasks and utilizes residual connections with attention mechanisms to identify complex patterns in tabular data. For this study, we used a balanced version of the Diabetes Health Indicators dataset, which contains an equal number of both non-diabetic and diabetic cases constructed from the 2021 BRFSS dataset.

### A. Dataset and Experimental Setup

In this study, we utilize a publicly available diabetes dataset [8], [10] derived from the 2021 Behavioral Risk Factor Surveillance System (BRFSS) dataset. The dataset contains tabular biomedical records, where each row represents a unique individual with an outcome label indicating the presence (1) or absence (0) of diabetes as determined through clinical diagnosis.

Following the pre-processing protocol outlined in the original ALTARN paper, we apply the following transformations. Mathematically, for *m* patient features and *n* individuals:

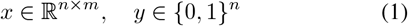

where *x* represents feature data and *y* the binary target. Binary features are mapped to 0 and 1:

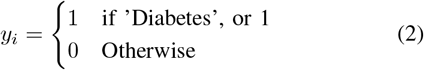

Non-numeric features are label encoded, mapping each unique category to an integer:

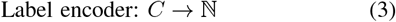

Missing values are imputed using the mean value of each respective feature computed on the training folds. All numeric features are standardized using z-score normalization:

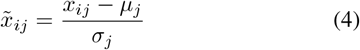

where *µ*_*j*_ is the mean and *σ*_*j*_ is the sample standard deviation for feature *j*.

### B. ALTARN Architecture Overview

The ALTARN model consists of the following components. The architecture begins by projecting the standardized feature vector into a latent representation of dimension 128 with batch normalization and ReLU activation:

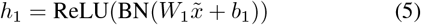

A sigmoid-based attention mechanism computes feature-wise weights:

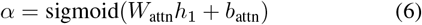

Element-wise multiplication produces the attention-weighted input:

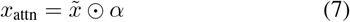

The residual block employs two parallel paths outputting dimension 64, combining a non-linear branch with dropout and a linear projection:

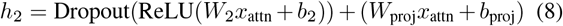

Further processing through a dense layer (dimension 32) with dropout and ReLU:

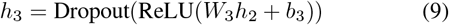

A final sigmoid activation produces the predicted probability *ŷ* of diabetes:

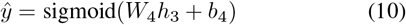

### C. Training Configuration

Following the original ALTARN implementation, we train the model using the Adam optimizer with a learning rate of 10^−3^ and binary cross-entropy loss. Training proceeds for up to 200 epochs with a batch size of 32. Early stopping with a patience of 10 epochs monitors validation loss and restores the best weights. Dropout regularization uses a rate of 0.2.

### D. Evaluation Protocol

To ensure robust evaluation, we employ 5-fold stratified cross-validation, maintaining the proportion of positive and negative instances across all folds. For each fold, the model is trained from scratch on four folds and validated on the remaining fold. Z-score normalization parameters are fitted solely on the training data and applied to the validation data to prevent data leakage. All random operations are seeded (seed=42) to ensure reproducibility.

**Fig. 1.**
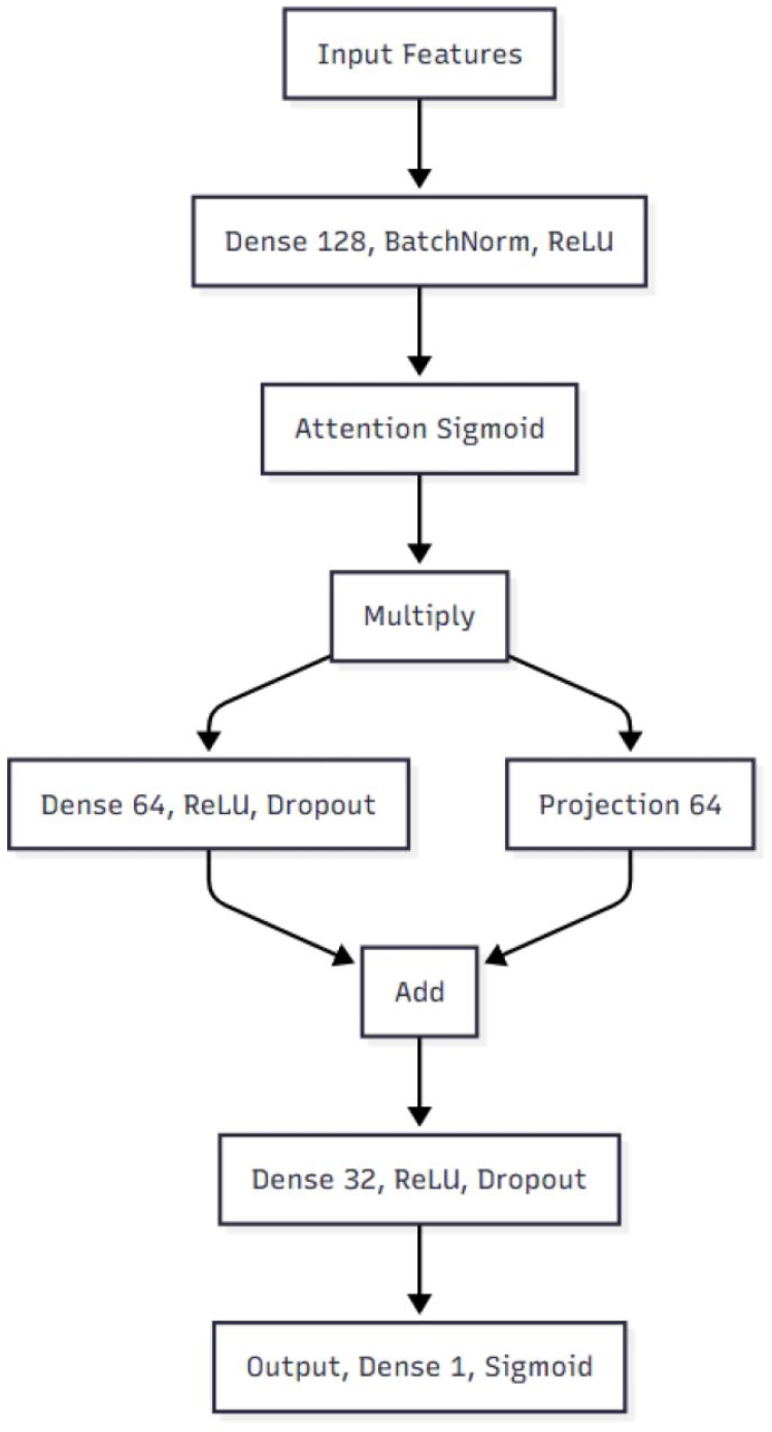
ALTARN Architecture

## III. RESULTS AND ANALYSIS

As shown in Fig. 2, ALTARN achieved an average training accuracy of 75.22 and a validation accuracy of 74.61. The model attains a mean validation precision of 72.36, a mean validation recall of 79.69, and a mean F1 score of 75.83. These results indicate a reasonably balanced trade-off between correctly identifying diabetic cases and limiting false positives on the balanced BRFSS 2021 diabetes indicators dataset.

**Fig. 2.**
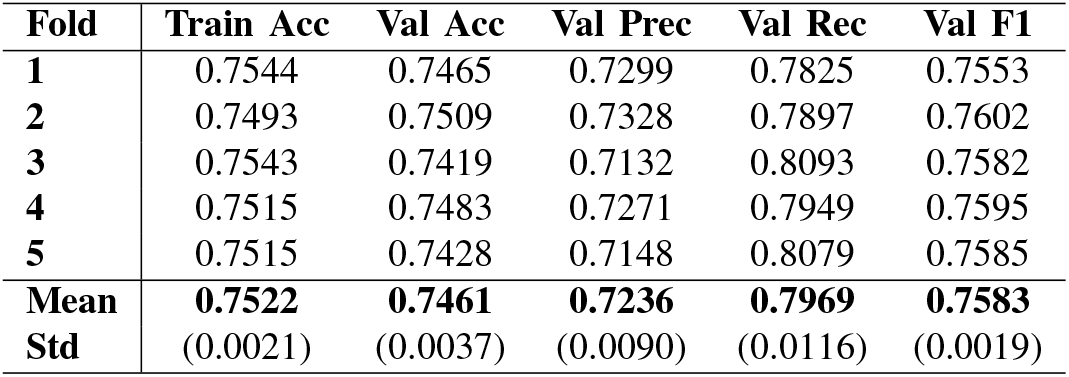
5-fold stratified cross-validation results of ALTARN on balanced BRFSS 2021 Diabetes Health Indicators data. Metrics are *mean* ± *std*.

Based on Fig. 3, the algorithm predicted 4549 instances in Healthy where they were Truly Healthy (True Negatives), and 5424 instances in Diabetes when it was Truly Diabetes (True Positives). However, the ALTARN incorrectly predicted 1290 cases in Healthy when they were Truly Diabetes (False Negatives), and 2164 cases in Diabetes when they were Truly Healthy (False Positives). The algorithm achieves a balanced tradeoff between recall and accuracy with relatively few false negatives compared to false positives, indicating that most diabetic cases were correctly identified.

**Fig. 3.**
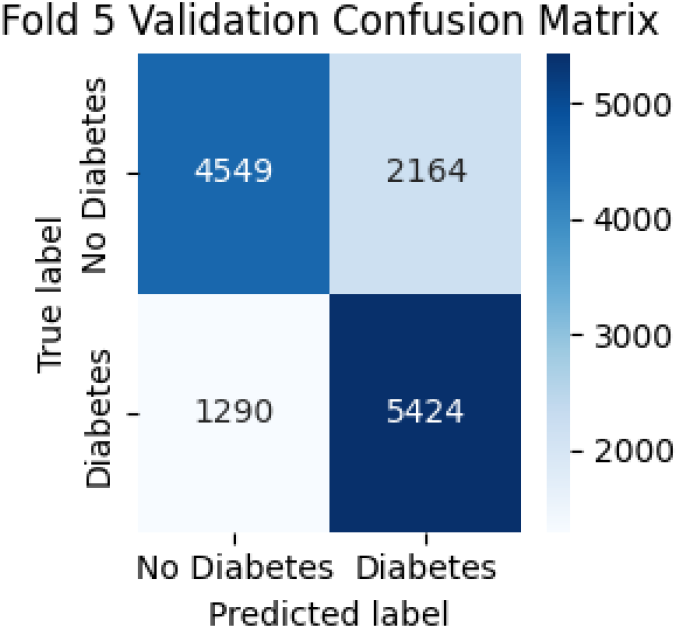
5-fold cross-validation confusion matrices for ALTARN.

**Fig. 4.**
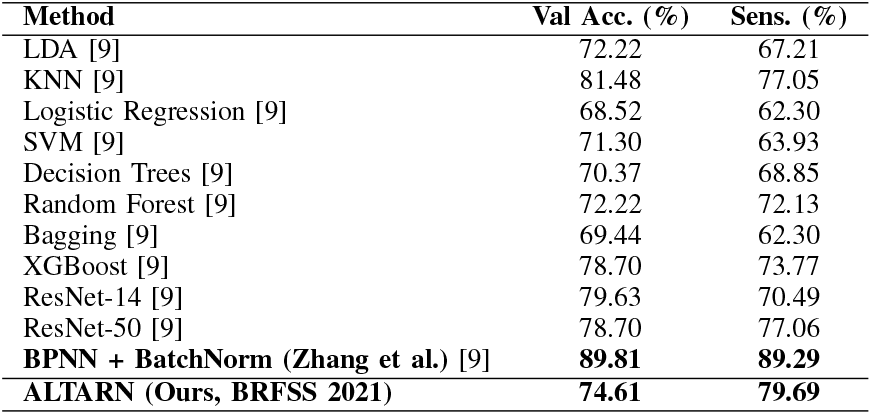
Benchmark comparison between deep learning models for diabetes diagnosis. Results for BPNN + BatchNorm and classical baselines are taken from Zhang et al. [9] on the Pima Indians Diabetes dataset, while ALTARN results are obtained on the balanced BRFSS 2021 Diabetes Health Indicators dataset.

In order to benchmark ALTARN, the model was compared against other varying classical and deep learning approaches, from Decision Trees to Support Vector Machines (SVMs) from a 2024 study. Although Zhang et al.’s BPNN with batch normalization has the highest sensitivity on the PIMA Indians dataset, ALTARN achieves strong sensitivity on a much larger, balanced 2021 BRFSS cohort, which highlights that an attention-based residual architecture generalizes well to real-world population-level health indicators.

## IV. Discussion

ALTARN, an attention-based residual neural network, to DM classification on the BRFSS 2021 dataset. The model achieved a validation accuracy of 74.61% with notably high recall (79.69%), which indicates strong performance in the identification of diabetic cases. Indeed, the high sensitivity provides clinical value as false negatives in diabetes screening can delay critical interventions and lead to severe complications.

ALTARN provides advantages over standard deep learning approaches: the sigmoid-based attention weights identify which features contribute most to classification decisions and diabetes risk factors at the population level emerge. The confusion matrix analysis revealed 1290 false negatives compared to 2164 false positives. In diabetes screening missed diabetic case often carries greater consequences than a false alarm. However, the precision of 72.36% indicates room for improvement in the reduction of false positives, which could decrease unnecessary follow-up tests and associated healthcare costs.

Comparison with benchmark models reveals important dataset-dependent performance patterns. Zhang et al.’s BPNN achieved 89.81% accuracy on the PIMA Indians dataset [9], [11], substantially higher than ALTARN’s 74.61% on BRFSS 2021. However, direct comparison is problematic due to fundamental dataset differences. PIMA Indians contains 768 samples with 8 features, while BRFSS 2021 contains over 250,000 samples with 21 features. The larger, more heterogeneous BRFSS cohort likely contains greater noise and population diversity, which naturally leads to lower accuracy but better generalization to real-world screening scenarios.

The consistent performance across all five cross-validation folds, with standard deviations below 1.2% for all metrics, confirms model stability. Albeit, some limitations are present:first, the balanced dataset does not reflect real-world diabetes prevalence (approximately 10 to 15%), which may affect model calibration in clinical settings. Secondly, the binary classification framework does not distinguish between Type 1 and Type 2 diabetes or prediabetic states, which limits clinical utility. Despite these limitations, the moderate accuracy suggests that tabular health indicator data alone may be suffiecient to help in diabetes prediction.

## V. Conclution

ALTARN, a tabular attention-based residual neural network, to binary classification of DM which achieved a mean validation accuracy of 74.61%. When combined with residual connections capture complex patterns in large-scale, population-level health data, ALTARN’s performance on BRFSS 2021 is modest compared to specialized models on smaller benchmark datasets like PIMA Indians. ALTARN exhibits robust performance on a substantially larger and more diverse real-world cohort. Future work should explore ensemble approaches that combine ALTARN with other architectures, analyze attention weights to identify key diabetes risk factors, and extend the model to multi-class classification to distinguish between Type 1 and Type 2 diabetes.

## Data Availability

All data produced in the present study are available upon reasonable request to the authors.

